# Molecular, Transcriptomic, and Proteomic Characterization of *Plasmodium* Infections that Evade Detection by Rapid Diagnostic Tests in Mizan Aman, Ethiopia

**DOI:** 10.64898/2026.01.04.25343089

**Authors:** Maria Nikulkova, Anne Kessler, Ziyi Wang, Abhishek Patel, Tirusew Tolessa, Taye Teka, Daniel Tesfaye, Biniam Lukas, Deje Lemessa, Marta Zemede, Fikirte Legesse, Harsh Srivastava, Steven A. Sullivan, Guiyun Yan, Delenasaw Yewhalaw, Jane M. Carlton

## Abstract

**Background:** Failure of rapid diagnostic tests (RDTs) to detect *Plasmodium* parasites in peripheral blood of individuals is a major barrier to successful case management and control of malaria in Ethiopia. Characterizing factors contributing to RDT failure is essential if malaria control and elimination strategies are to succeed.

**Methods:** We consented and enrolled 148 individuals with suspected malaria presenting to health clinics in Mizan Aman, Ethiopia. We administered a clinical questionnaire, diagnosed the presence of malaria parasites via RDT, and collected venous blood. Samples were assayed using molecular methods to detect parasite DNA, *Plasmodium* species, parasite load, and *pfhrp2* and *pfhrp3* gene deletions. RNA-seq libraries and LC-MS proteomics data were generated from all molecularly confirmed *P. falciparum*-infected individuals.

**Results:** We identified 29/148 (27.9%) individuals as *P. falciparum* PCR positive with 26/29 (89.7%) false negative by a P.f/Pan RDT. RDT+ *P. falciparum* and *P. vivax* infections had higher parasite densities than RDT- infections. Of the 29 *P. falciparum* infections, 27 (93.1%) had deletions in both *pfhrp2* and *pfhrp3* genes, and 22 (75.9%) had negligible *pfhrp2* transcripts. Ten *P. falciparum* samples had detectable PfLDH peptides, but no samples had PfHRP2 or PfHRP3 peptides detectable by LC-MS.

**Conclusions:** Our molecular, transcriptomic, and proteomic characterization of *P. falciparum* infections that fail detection by PfHRP2/pLDH-based RDTs in Mizan Aman, Ethiopia, revealed a heterogeneous array of factors that could be responsible for the observed RDT failure.

---

Rapid diagnostic tests (RDTs) are a key tool for detecting *Plasmodium* parasites in peripheral blood of individuals presenting at health facilities in malaria-endemic countries. RDTs are immunochromatographic assays: when a drop of infected blood is applied, parasite antigen-human antibody complexes are formed that migrate along a nitrocellulose membrane, creating a visible line if the target parasite antigen is present [1]. The majority of commercially available RDTs specific for *Plasmodium falciparum* target histidine-rich protein 2 (PfHRP2), a highly abundant antigen in the blood of infected individuals [2]. Its less abundant homologue histidine-rich protein 3 (PfHRP3) provides a complementary detection pathway when PfHRP2 levels are low. *Plasmodium* lactate dehydrogenase (pLDH)-based RDTs [3] detect the distinct isoforms of pLDH enzyme produced by other human malaria parasite species, i.e., *P. vivax, P. ovale*, and *P. malariae*. RDTs provide rapid point-of-care diagnoses in resource-limited areas but have limitations including insensitivity to low parasitemias [4, 5] and insufficient specificity to reliably distinguish *Plasmodium* species [6, 7].

Especially concerning is the spread of *P. falciparum* strains with partial or full deletion of the *pfhrp2* and *pfhrp3* genes [8–10], resulting in “diagnostic escape”, i.e., missed or misdiagnosed cases of *P. falciparum*.

Ethiopia has experienced a significant increase in the estimated number of malaria cases in recent years, reaching 6.9 million over a five-year period (2019-2023), and an 82% increase in incidence between 2022-2023 [11]. The country set a nationwide malaria elimination goal of 2030 and implemented use of RDTs for early diagnosis of *Plasmodium* infections as part of its National Malaria Elimination Strategy Plan (NMSP) [12]. The spread of *P. falciparum* strains carrying deletions of the *pfhrp2/3* genes in Ethiopia [13–16] led the World Health Organization to urge greater surveillance of such strains [17].

Here we report characterization of the *Plasmodium* infection status of 148 participants who presented at health clinics in October 2022 in Mizan Aman, Ethiopia, and molecular, transcriptomic, and proteomic investigations of 29 *P. falciparum* infections that evaded RDT detection. Our finding of an overwhelming diagnostic failure of the P.f/Pan RDT used and the significant heterogeneity in sample characteristics that could be responsible for RDT failure highlights the urgent need to look for better parasite biomarkers for the development of improved RDTs.

## METHODS

### Study Sites and Participant Enrollment

Participants were enrolled in October 2022 from Mizan Teaching Hospital, Mizan Health Center, and Kite Health Center in Mizan Aman, southwestern Ethiopia, a lowland area with *P. falciparum* and *P. vivax* where the major malaria peak occurs after the rainy season in September-December [18, 19]. The study protocols were approved by the National Research and Ethics Review Committee (NRERC), Ministry of Education, and the Federal Democratic Republic of Ethiopia (Reference No. 03/246/9S/22). Informed consent/assent was obtained prior to study enrollment. A clinical questionnaire was administered, axillary body temperature measured by a physician for participants, and fever defined as a temperature ≥37.3°C. Participants self-reported whether they had been diagnosed with malaria in the previous year.

### Sample Collection and RDT Classification

Approximately 5 µL of finger-prick blood from each participant was applied to a Bioline™ Malaria Ag P.f/Pan (Abbott) RDT which detects *P. falciparum* HRP2 antigen (P.f line) and *Plasmodium* LDH antigen (Pan line) in human whole blood. A positive control line indicates a valid test. All samples tested were classified according to the manufacturer’s guide: Pan+P.f- indicates non-falciparum malaria (P.v, P.m, P.o); Pan+P.f+ indicates either a *P. falciparum* infection or a mixed species infection where *P. falciparum* is present alongside one or more other *Plasmodium* species (*e.g.*, P.v, P.m, P.o); and Pan-P.f- indicates a negative RDT result (Abbott quick reference guide).

Two vacutainers of venous whole blood were collected from each participant: 7.5 mL in acid-citrate-dextrose glass collection tubes (ACD; Becton, Dickinson and Company) and 3 mL in Tempus^TM^ Blood RNA Tube (Applied Biosystems). The ACD vacutainer blood samples were transferred to 15 mL falcon tubes, centrifuged at 4000 rpm for five minutes, and the plasma supernatant pipetted into separate cryovials. Samples were stored at -80°C until transfer to New York University in January 2023 for further processing.

### DNA and RNA Extraction

Total genomic DNA was extracted from the ACD vacutainers using the Qiagen® Blood and Cell Culture Midi Kits, and tested for quality and concentration using a Qubit fluorometer. Total RNA was extracted from Tempus^TM^ tubes using the Tempus™ Blood RNA Systems kit (Thermo Fisher). RNA quality control was undertaken using Agilent RNA ScreenTape System and Qubit instruments to determine RNA Integrity Number and total RNA concentrations.

### *Plasmodium* Species-Specific PCR

Samples were tested for (1) *P. falciparum* and *P. vivax* parasite DNA using a single step, multiplexed PCR assay adapted from Demas *et al*. 2011 [20] that targets the multicopy loci Pfr364 (*P. falciparum*) and Pvr47 (*P. vivax*); and (2) *P. ovale* and *P. malariae* DNA using a nested, multiplexed PCR targeting the 18S rRNA gene [21].

### Quantification of Parasitemia by 18S rRNA qPCR Assay

The relative parasitemias for *P. falciparum*, *P. vivax*, and mixed *P. falciparum*/*P. vivax* infections were determined using qPCR of asexual parasite 18S rRNA gene through modification (using DNA instead of RNA) of a published quantitative reverse transcription PCR assay [22, 23]. *P. falciparum* 3D7 was grown *in vitro* and a parasite dilution series undertaken as described [24]. Ten sets of culture dilutions were run in triplicate in a 96-well qPCR plate on a Roche LightCycler 480 II to estimate average Ct values and create a dilution series curve based on 18S rRNA gene copy number. A cutoff of 35 cycles was used to define *P. falciparum* positive samples. Amplification of the single-copy human actin gene from human DNA (obtained from Promega and Novagen), *P. falciparum* 3D7 (DNA nos. 533 and 107)*, P. vivax* (IndiaNYC DNA no. 538, IndiaVII DNA no. 220), and a plasmid containing the *P. vivax* 18S rRNA (MRA 178 from BEI Resources, contributed by Peter A. Zimmerman) were used to normalize the qPCR, adjust for noise, and set the number of cycles required to cross the threshold level of fluorescence.

Statistical analysis of 18S rRNA gene expression (as an estimate of parasitemia) and RDT (Pan-P.f-, Pan+P.f-, and Pan+P.f+) result was performed using pairwise Wilcoxon rank-sum tests. The Benjamini-Hochberg (BH) correction for multiple testing (adjusted p <0.05) was used for PCR-confirmed *P. falciparum* samples, but no BH adjustment was used for PCR-confirmed *P. vivax* samples as there were only two RDT classifications (Pan-P.f- and Pan+P.f-). Relationships between continuous variables (*i.e.,* parasitemia) were assessed using Spearman’s correlation with a *p*-value < 0.05 considered statistically significant; a Shapiro-Wilks test was performed to determine if the data were normally distributed; and relative parasitemia visualized on QQ-plots. Sample MTH-018 (Pan-P.f-) was excluded from analysis since it did not amplify during qPCR (reported as Ct = 0 by instrument software) but showed a band during ssPCR amplification.

### *pfhrp2*/*3* Deletion Assay and Analysis

A *pfhrp2* and *pfhrp3* deletion assay adapted from Grignard *et al.* 2020 [25] was used on all PCR-confirmed *P. falciparum* (n=29) and coinfected (n=13) samples, in triplicate in 96-well plates with 5 µL DNA template or control. Fluorescent probe markers were modified to ensure compatibility with the Roche LightCycler 480 II used (**Supplementary Table 1**). Each assay included positive controls of two human DNA samples, three *P. falciparum* 3D7 (*pfhrp2*+/*pfhrp*3+) samples, one *P. falciparum* Dd2 (*pfhrp2*-/*pfhrp3*+) sample, and three nuclease-free water non-template negative controls. Samples were also tested using the *pfldh* and human beta tubulin (*HumTuBB*) genes [25] for sample quality control.

### RNA-seq Library Preparation, Sequencing, and Analysis

Total RNA from the 29 *P. falciparum* samples was used as input into an RNA HyperPrep Kit with RiboErase and Globin Depletion (KAPA) to generate cDNA libraries for paired-end RNA-seq. Samples were uniquely barcoded using KAPA Plate UDI Primer mix using a reaction of 25 µL KAPA HiFi HotStart ReadyMix (2x) and 5 µL UDI Primer following the KAPA RNAHyperCap Workflowv1.1 protocol. Amplification occurred for 6 minutes at 94°C during the fragmentation, priming, and eluting step, and the final thermocycling profile was run with 10 cycles of amplification. cDNA libraries were quality checked on a TapeStation D1000 HS, multiplexed, and sequenced on an Illumina NovaSeq 6000 instrument (NYU Genomics Core) to generate 2 x 100 bp reads (insert size 200-300 bp).

RNA-seq reads underwent quality control and preprocessing using FastQC, and Trimmomatic for trimming of adapter and lowquality score nucleotides, and FastQC to visualize the quality matrix before and after trimming. Processed RNA-seq reads were mapped to the human genome (Ensembl build GRCh38) using STAR [26]. Unmapped reads were collected and aligned to the *P. falciparum* 3D7 genome sequence using STAR. Aligned BAM files and BAM indexes were imported into Integrative Genomics Viewer [27] and visualized against the *P. falciparum* 3D7 genome sequence region containing *pfhrp2* (PF3D7_0831800, chromosome 8), *pfhrp3* (PF3D7_1372200, chromosome 13), and *pfldh* (PF3D7_1324900, chromosome 13). DESeq2 [28] was used to compare expression levels across these three genes and to normalize raw counts for sequencing depth, making them comparable across samples.

### Plasma Proteomics by LC-MS and Data Analysis

Plasma aliquots from the 29 *P. falciparum*-infected subjects were analyzed by Liquid Chromatography-Mass Spectrometry (LC-MS) using Seer Inc., Proteograph Assay utilizing nanoparticles to enrich proteins across the dynamic range of the plasma proteome. Following nanoparticle binding, enriched proteins were subjected to on-particle trypsin digestion. The resulting peptide pools were desalted, prepared for LC-MS analysis using a Thermo Scientific Ultimate 3000 UHPLC system with a 20-minute chromatographic gradient, coupled to an Orbitrap Astral mass spectrometer operated in data-independent acquisition (DIA) mode with a 24-minute acquisition cycle. Peptide data were analyzed using DIA-NN [29] and the dataset searched against combined human (taxonomy ID: 9606) and *P. falciparum* (taxonomy ID: 36329) Interpro databases [30]. Trypsin specificity allowed for up to two missed cleavages, with methionine oxidation as a variable modification, cysteine carbamidomethylation as a fixed modification, and a 1% FDR threshold. Identified proteins, peptides, and intensity values were cross-checked for the presence of PfHRP2 (PF3D7_0831800), PfHRP3 (PF3D7_1372200) and PfLDH (PF3D7_1324900) based on their unique identifiers.

*In silico* trypsin digestion was simulated using Rapid Peptide Generator (RPG) and set to cleave at lysine (K) and arginine (R) residues unless followed by proline (P). Theoretical peptides between 6–30 amino acids and 500–3000 Da were used to calculate sequence coverage as described [31]. Proteolytic digestion of the entire *P. falciparum* proteome was undertaken using 46 enzymes available in RPG. Coverage rates were compared to identify those enzymes that were optimal for detecting PfHRP2, PfHRP3, and PfLDH. The amino acid compositions of PfHRP2, PfHRP3, and PfLDH protein sequences were calculated and compared to the average composition of all *P. falciparum* proteins in the LC-MS dataset.

## RESULTS

### Patient Characteristics

We enrolled 148 study participants over three days in October 2022 at three health facilities in Mizan Aman town, Ethiopia, as part of a pilot study utilizing multi-omics methods to investigate *Plasmodium-*host interactions. Of the 148 participants, 59% (n=87) were female, 58% (n=86) were aged 11-25, and most participants were of Bench ethnicity (68.2%; n = 101) (**Table 1A**). Half of the participants (50%; n=74) reported a malaria diagnosis within the past year (**Table 1B**). Fifty-eight (39.2%) participants were febrile at the time of enrollment. The majority of participants (56.8%; n=84) reported fever with at least one other symptom (*e.g.,* abdominal pain, chills/shivering, coughing, diarrhea, difficulty breathing, fatigue headache, joint pain, loss of appetite, malaise, muscle pain, vomiting) within two days of coming to the clinic (**Table 1B**).

**Table 1.** Socio-demographic and clinical characteristics of study participants (n=148) . **ALT Text:** Table summarizing the socio-demographic and clinical characteristics of 148 study participants. Includes sex, age, ethnicity, occupation, education, self-reported previous malaria diagnosis, and symptoms.

| Study variable | Category | n | % |
| --- | --- | --- | --- |
| **A. Sociodemographic characteristics** |  |  |  |
| **Sex** | Male | 61 | 41.2 |
|  | Female | 87 | 58.8 |
| **Age Strata (Years)** | 4-10 | 9 | 6.1 |
|  | 11-18 | 39 | 26.4 |
|  | 19-25 | 47 | 31.8 |
|  | 26-35 | 30 | 20.3 |
|  | 36-45 | 15 | 10.1 |
|  | 46+ | 8 | 5.4 |
| **Ethnicity** | Amhara | 23 | 15.5 |
|  | Bench | 101 | 68.2 |
|  | Kafficho | 11 | 7.4 |
|  | Oromo | 6 | 4.1 |
|  | Multiple Ethnicities | 2 | 1.4 |
|  | Other Ethnicity | 5 | 3.4 |
| **Occupation** | Agricultural Worker | 27 | 18.2 |
|  | Housewife | 19 | 12.8 |
|  | Office Worker/Teacher | 8 | 5.4 |
|  | Own Business/Shopkeeper | 3 | 2.0 |
|  | Student | 50 | 33.8 |
|  | Trader | 9 | 6.1 |
|  | Unemployed | 5 | 3.4 |
|  | Other | 3 | 2.0 |
|  | Not Reported | 24 | 16.2 |
| **Education** | Illiteracy/Never attend school | 18 | 12.2 |
|  | Preschool (<5 years) | 3 | 2.0 |
|  | Primary School | 47 | 31.8 |
|  | Middle (lower 2nd) School | 21 | 14.2 |
|  | High (upper 2nd) School | 6 | 4.1 |
|  | University/Higher Education | 9 | 6.1 |
|  | Not Reported | 44 | 29.7 |

| **B. Clinical characteristics** |  |  |  |
| --- | --- | --- | --- |
| Axillary Temperature (°C) | ≤36.5 | 17 | 11.5 |
|  | 36.6-37.2 | 33 | 22.3 |
|  | ≥37.3 (fever) | 58 | 39.2 |
|  | Not Reported | 40 | 27.03 |
| Previous Malaria Diagnosis | Yes | 74 | 50 |
|  | No | 28 | 18.9 |
|  | Not Reported | 46 | 31.1 |
| Symptoms in the last 2 days | Yes | 84 | 56.8 |
|  | No | 64 | 43.2 |

### Detection of *Plasmodium* Parasites by RDT and Species-Specific PCR

We diagnosed 74/148 (50.0%) of the participants as Pan+P.f- (*i.e.,* non-*falciparum Plasmodium* infection), 4/148 (2.7%) as Pan+P.f+ (*i.e., P. falciparum* single or mixed species infection), and 70/148 (47.3%) as Pan-P.f- (*i.e.,* negative for *Plasmodium* infection) by RDT. None of the participants were Pan-P.f+ by RDT. Participants identified as Pan+P.f- and Pan+P.f+ were characterized as “RDT+”, and participants identified as Pan-P.f- were classified as “RDT-”.

We confirmed *Plasmodium* infection status by species-specific PCR. Of the 148 participant samples, 70% (n=104) were *Plasmodium* positive by ssPCR, of which 59.6% (n=62) were *P. vivax*, 27.9% (n=29) were *P. falciparum*, and 12.5% (n=13) were *P. vivax/P. falciparum* co-infections (**Figure 1**). The remaining 29.7% (n=44) samples were uninfected, and no samples were positive for *P. malariae* or *P. ovale*.

**Figure 1.**
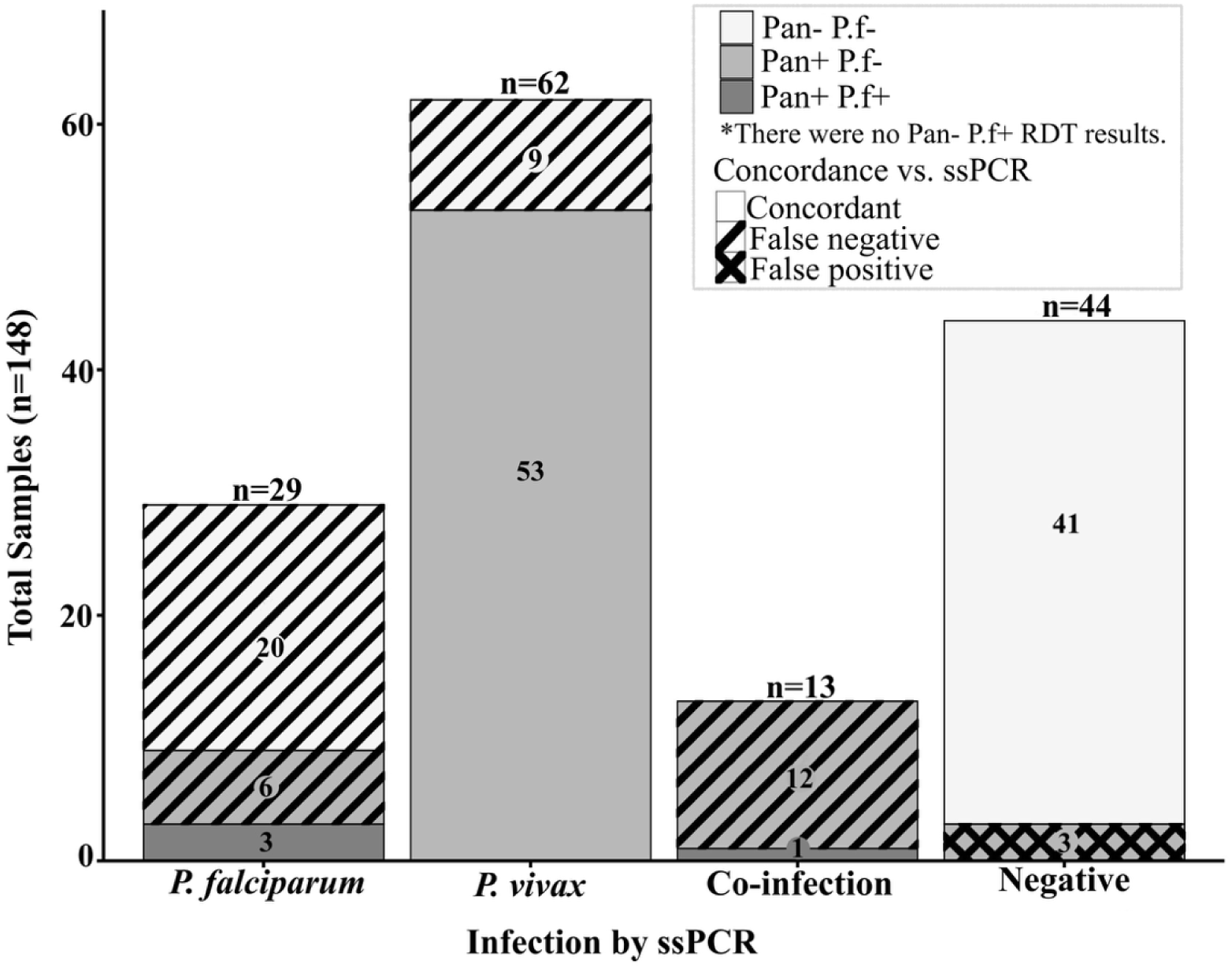
***Plasmodium* infection status by ssPCR and RDT**. Columns indicate the number of subjects (y-axis) with ssPCR results for each of the four infection categories (x-axis). Each column is colored according to the participant RDT results, and concordance between the RDT result and ssPCR is also shown (see key). Abbreviations: RDT, rapid diagnostic test; ssPCR, species-specific PCR. **ALT Text:** Stacked bar chart showing total study subjects separated by infection category based on species-specific PCR results. Infection categories include *P. falciparum*, *P. vivax*, Co-infections (*P. falciparum* and *P. vivax*), and negative. Bars are colored by RDT results, and concordance between diagnostic methods is also depicted.

Of the 62 participants *P. vivax* positive by ssPCR, 53/62 (85.5%) were accurately classified by RDT (*i.e.,* Pan+P.f-) while the remaining 9/62 (14.5%) *P. vivax* infections were false negatives by RDT (*i.e.,* Pan-P.f-) (**Figure 1**). Of the 29 *P. falciparum* ssPCR positive samples, 3/29 (10.3%) wer accurately classified by RDT (Pan+P.f+), while the majority 26/29 (89.7%) were false negative by RDT (*i.e.,* six Pan+P.f- and 20 Pan-P.f-). Of the 13 PCR confirmed *P. vivax/P. falciparum* co-infections, only one was accurately identified by RDT (Pan+P.f+). The remaining 12 co-infected samples were false negatives by RDT (Pan+P.f-) (**Figure 1**).

Two co-infected samples (MTH-009 and MTH-031) were *P. vivax* positive and *P. falciparum* negative by qPCR despite amplification of both *P. vivax* and *P. falciparum* targets by ssPCR. A total of 44 samples were negative for *Plasmodium* infection by ssPCR. Of these confirmed negatives, 3/44 (6.8%) were false positives by RDT (*i.e.,* Pan+P.f-) while the remaining 41/44 (93.2%) were true negatives (Pan-P.f-) (**Figure 1**).

### Contribution of low parasitemic infections to observed RDT failure

To assess whether low-density infections contributed to the observed RDT misclassifications, we estimated relative parasitemia by a qPCR assay targeting the 18S rRNA gene. Of the ssPCR-confirmed *P. falciparum* samples (n=29; 3 Pan+P.f+, 6 Pan+P.f- and 20 Pan-P.f-), the Pan-P.f- samples (*i.e.,* RDT-) had lower parasite densities when Ct values were plotted against log_10_ *P. falciparum* 18S rRNA gene copies in a dilution series (**Figure 2A**). We found a statistically significant difference (adjusted *p*=0.006) in parasite density between the Pan-P.f- (RDT-) and the Pan+P.f- (*i.e.,* RDT+ for non-*falciparum Plasmodium*). We found no statistically significant difference in parasite density between the Pan-P.f- (RDT-) and Pan+P.f+ (*i.e.,* RDT+ for *P. falciparum* mono or mixed species infection) groups (adjusted *p*=0.104), although the Pan+P.f+ samples had generally higher parasite densities that were not significantly different from the Pan+P.f- samples (adjusted *p* = 0.905) (**Figure 2A**).

**Figure 2.**
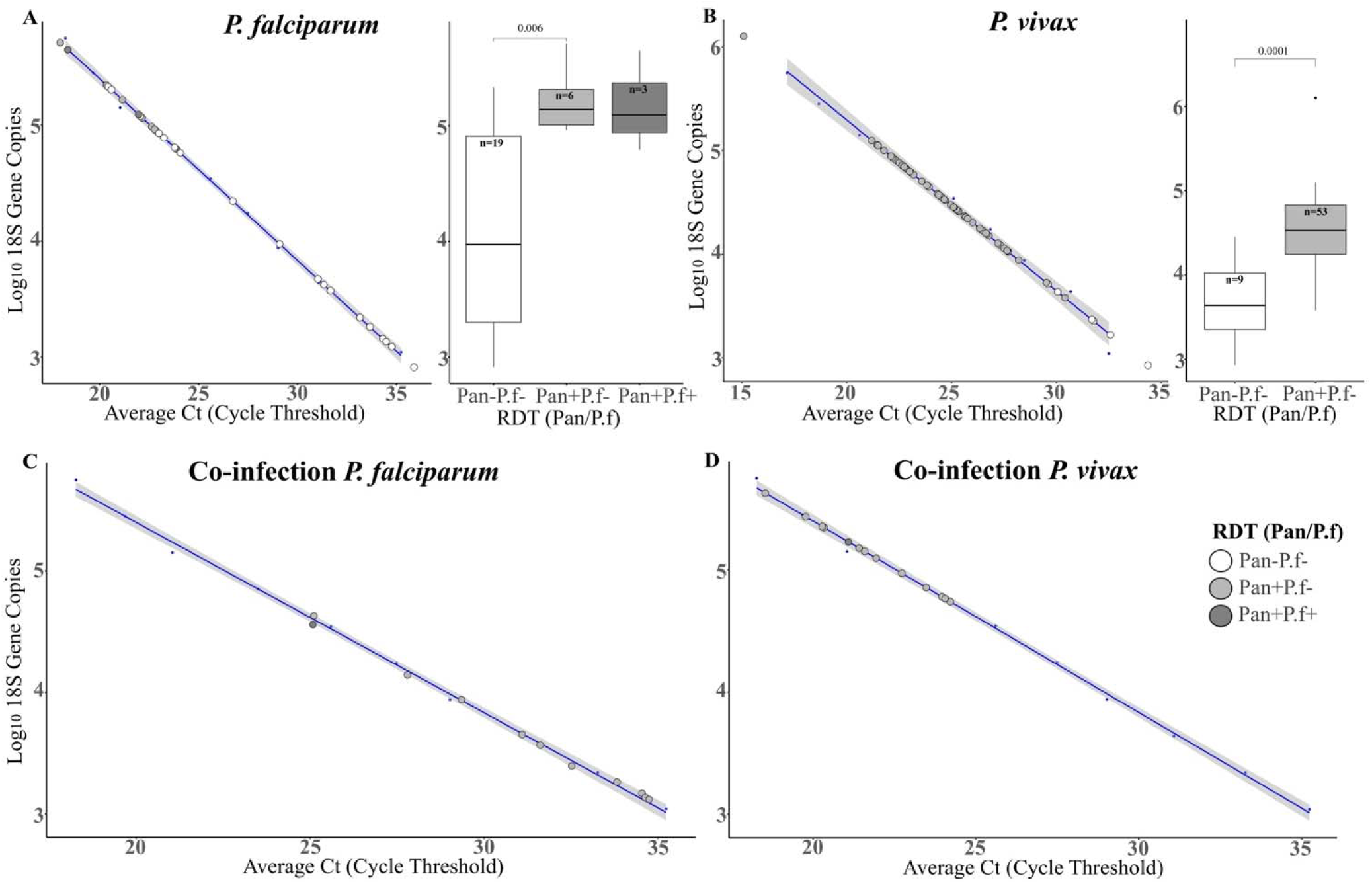
Parasitemia correlation and RDT result. **A**. Scatter plot of *P. falciparum* 18S rRNA gene copy number as a function of average Ct with trend line. Inset box plot depicts *P. falciparum* 18S rRNA gene copy number by Pan/P.f RDT result; outlier isolate MTH-018 was removed from the plot. **B**. Scatter plot of *P. vivax* 18S rRNA gene copy number as a function of average Ct with trend line. Inset box plot depicts *P. vivax* 18S rRNA gene copy number by Pan/P.f RDT result. **C-D.** Estimation of 18S rRNA gene copy number in co-infections, shown separately for *P. falciparum* (C; outliers MTH-009 and MTH-031 removed from the plot) and *P. vivax* (D) with trend lines. Panel headings indicate molecular-confirmed infection status. Key for all scatterplots: open circles = Pan-P.f- RDT-isolates; light grey circles = Pan+P.f- RDT+ isolates; dark grey circles = Pan+P.f+ RDT+. **ALT Text:** Three-panel figure showing the correlation of 18S rRNA gene copy number with parasitemia. Panel A: Scatter plot and boxplot of *P. falciparum* 18S rRNA gene copy number overall and by Pan/P.f RDT status, respectively. Panel B: Scatter plot and boxplot of *P. vivax* 18S rRNA gene copy number overall and by Pan/P.f RDT status, respectively. Panel C: Scatter plots for 18S rRNA gene copy number and RDT result for *P. falciparum* and *P. vivax* coinfections separated by species and colored by Pan/P.f RDT status.

For the ssPCR-confirmed *P. vivax* samples (n=62), higher parasitemias corresponded to Pan+P.f- (RDT+ for non-falciparum *Plasmodium*) samples compared to Pan-P.f- (RDT-) samples (**Figure 2B**; pairwise Wilcoxon rank-sum test, *p* = 0.0001). In mixed *P. vivax/P. falciparum* infections, species-specific Ct values (derived from distinct fluorescent markers for the two species) were higher for *P. falciparum* (**Figure 2C**) than *P. vivax* (**Figure 2D**) parasitemias. No co-infected samples were classified as Pan-P.f- (*i.e.,* RDT-). The single co-infected sample (MHC-031) that was accurately identified by RDT (*i.e.,* Pan+P.f+) also had the highest relative parasitemia for *P. falciparum* when compared to the 12 other co-infection samples (Pan+P.f-).

### Characterization of *pfhrp2* and *pfhrp3* deletions and expression further clarifies RDT results

*P. falciparum* isolates confirmed by ssPCR (n=29) were assayed for *pfhrp2* and *pfhrp3* gene deletions and the presence of *pfhrp2, pfhrp3*, and *pfldh* transcripts. First, the quality and integrity of parasite DNA was confirmed for the *P. falciparum* isolates by qPCR of *pfldh*, with 22 samples passing QC. Of those 22, 20 (90.9%) had deletions in both *pfhrp2* and *pfhrp3* (2 Pan+P.f+, 6 Pan+P.f-, and 12 Pan-P.f-) (**Table 2**). No isolates had deletions in only one of the two genes. Two isolates (MHC-003, Pan-P.f-and MHC-046, Pan+P.f+) with intact *pfhrp2* and *pfhrp3* genes had among the highest estimated *P. falciparum* parasitemias.

**Table 2.**
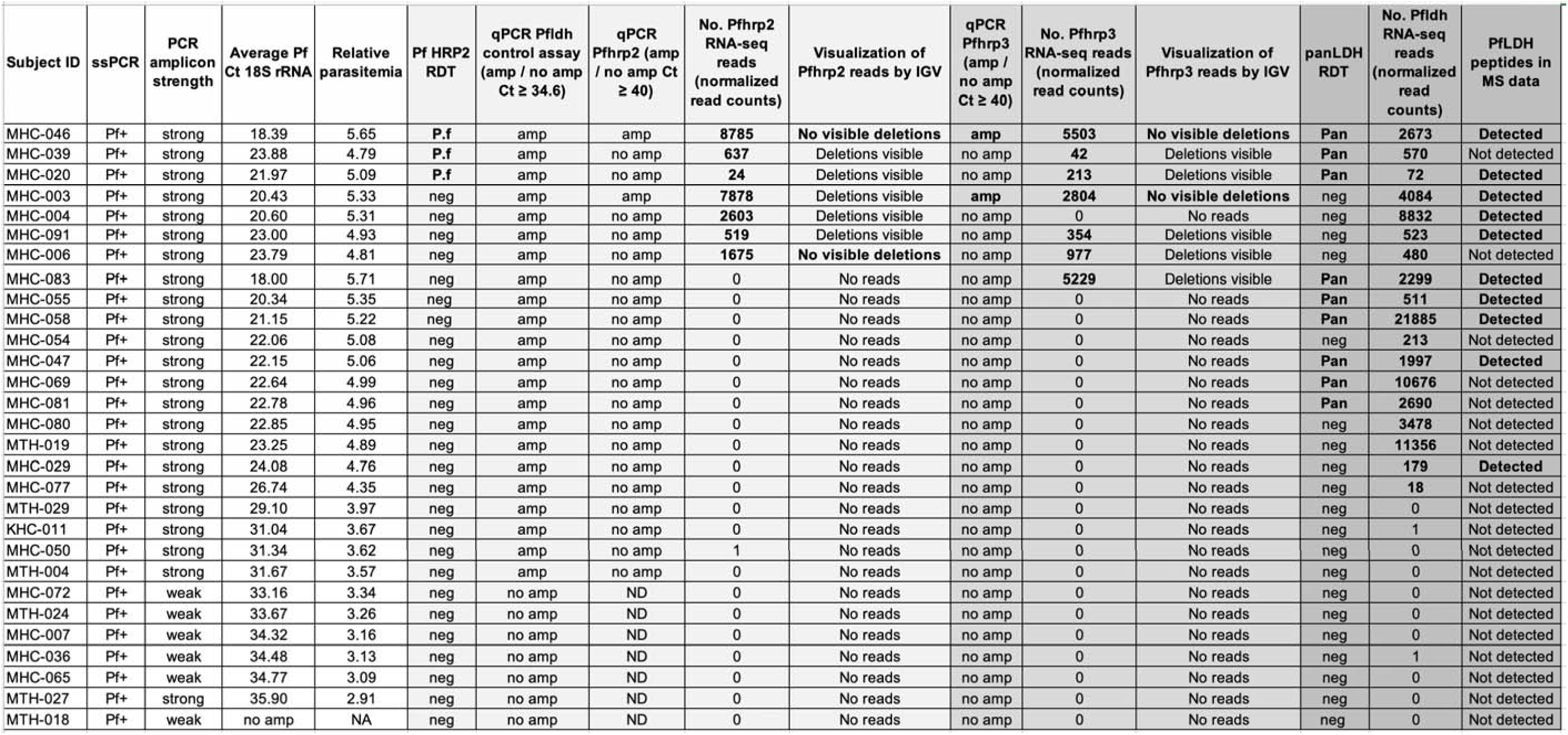
RDT, ssPCR, qPCR, RNA-seq, and proteomics assay results for PCR-confirmed *P. falciparum* subjects. . Isolates are ordered with the 3 RDT+ at the top, followed by isolates with *Pfhrp2* transcripts. Important results are in bold for ease of viewing, and assays for each of the three diagnostic genes/proteins of interest are grouped next to each other and shaded. Abbreviations: RDT, rapid diagnostic test; ssPCR, species-specific PCR; Ct, cycle threshold; amp, amplification; ND, not determined/missing data; IGV, Integrative Genomics Viewer; MS, mass spectrometry. **ALT Text:** Multi-column data table showing *P. falciparum* participants and their assay results across different methods.

RNA-seq data from the 29 ssPCR-confirmed *P. falciparum* isolates was used to assess *pfhrp2/pfhrp3* gene expression. Normalized read counts were determined for *pfhrp2*, *pfhrp3*, and *pfldh* loci and visualized. The two *P. falciparum* isolates with intact *pfhrp2* and *pfhrp3* genes (MHC-003 and MHC-046) had the highest number of mapped RNA-seq reads to *pfhrp2*. Eight isolates (3 Pan+P.f+, 5 Pan-P.f-) had >1 normalized read mapping to the *pfhrp2* locus, and seven isolates (3 Pan+P.f+, 1 Pan+P.f-, and 3 Pan-P.f-) had reads mapping to the *pfhrp3* locus. MHC-083 had no *pfhrp2* reads but had the highest number of reads for *pfhrp3* when compared against all isolates with visible deletions. The presence of *pfldh*-mapped reads was associated with higher parasitemia samples and those that amplified the *pfldh* control gene during the qPCR assay (**Table 2**).

### Proteomics of plasma from 29 *P. falciparum* isolates

We identified 1,221 unique peptides in plasma aliquots from the 29 *P. falciparum* infected subjects, and these mapped to 265 unique *P. falciparum* proteins. None of the peptides mapped to the PfHRP2 or PfHRP3 antigens. PfLDH peptides were identified in 10/29 *P. falciparum* plasma samples, comprising two Pan+P.f+ participants (MHC-020, MHC-046), four Pan+P.f- participants (MHC-047, MHC-055, MHC-058, MHC-083), and four Pan-P.f- participants (MHC-003, MHC-004, MHC-029, MHC-091). We undertook an *in silico* prediction of digestion with trypsin enzyme (the most widely used enzyme for MS) to investigate the unexpected absence of PfHRP2 and PfHRP3 in the proteomics dataset. This generated theoretical peptide sequences for PfHRP2, PfHRP3, and PfLDH within the LC-MS detectable range of 6–30 amino acids (**Table 3**). A lower number of peptides for PfHRP2 and PfHRP3 compared to PfLDH was predicted. Furthermore, the peptide sequence coverage for PfHRP2 (12.79%) and PfHRP3 (14.18%) was much lower than the 54.43% coverage observed for PfLDH and the average coverage of 62.61% determined for all *P. falciparum* proteins.

**Table 3.**
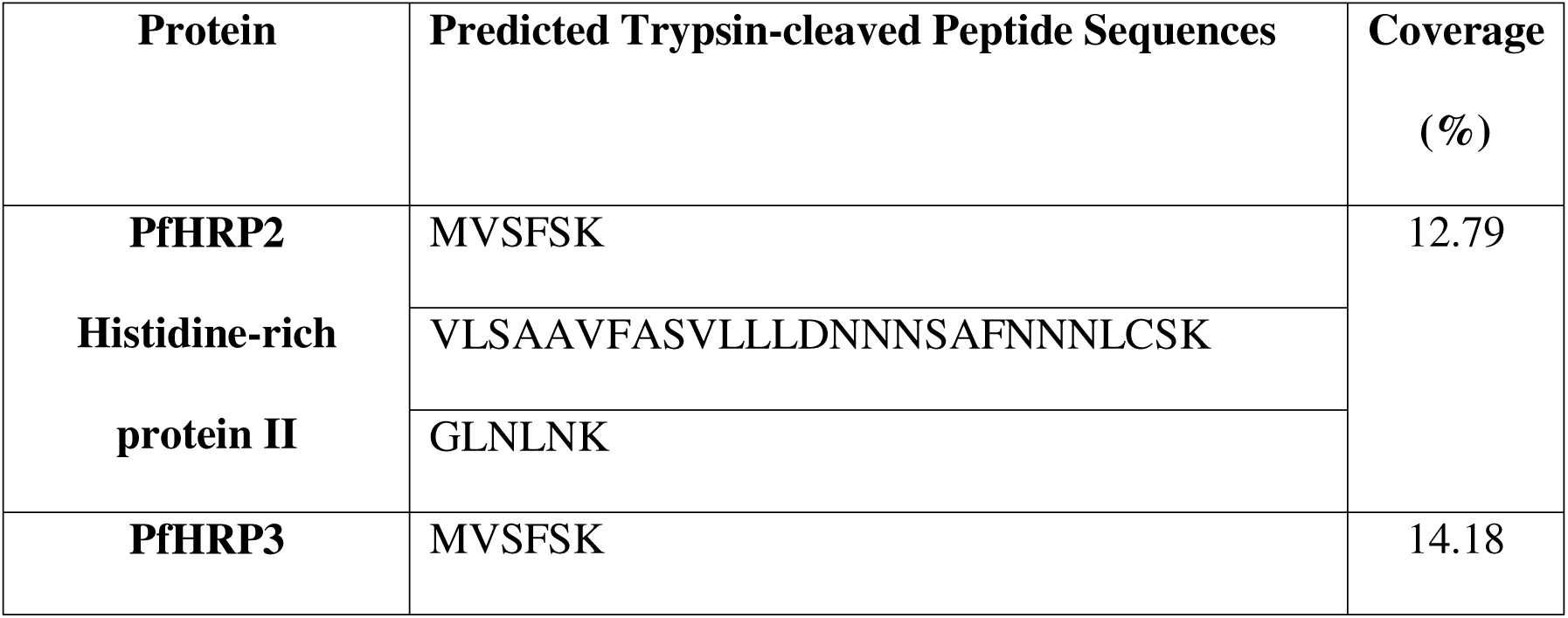
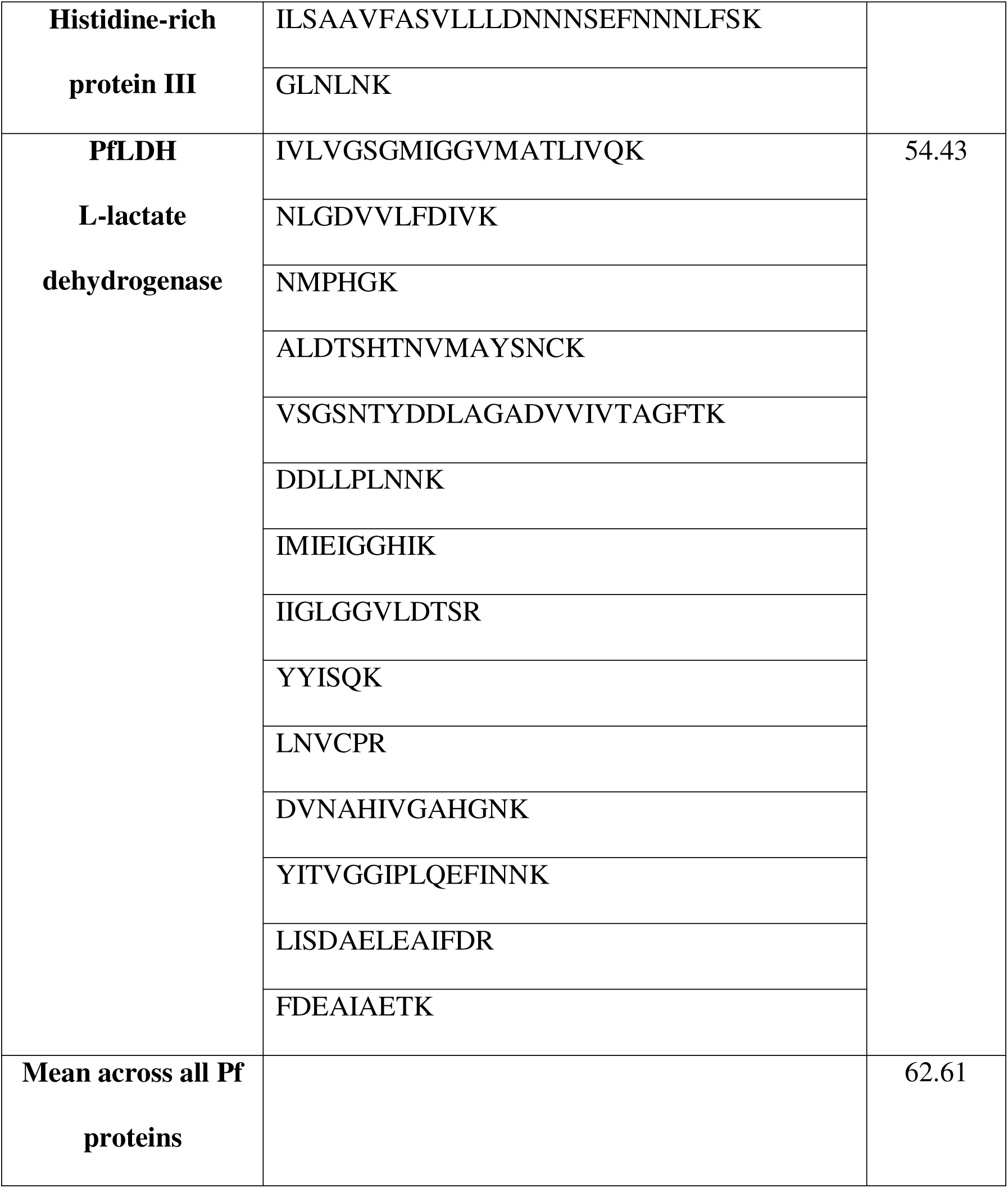
Coverage of *P. falciparum* proteins predicted from *in silico* trypsin digested peptides. **ALT Text:** Table listing predicted *P. falciparum* proteins. Includes coverage after trypsin digestion. Table includes protein name, trypsin-cleaved peptide details, and coverage percentage.

PfHRP2 and PfHRP3 are known to be histidine (H)-rich proteins. We analyzed the amino acid compositions of these genes and of the trypsin cleavage residues of arginine (R) and lysine (K); PfHRP2 (305 amino acids in length) has 2 arginine (R), 5 lysine (K), and 104 H residues; PfHRP3 (275AA) has 1 R, 6 K, and 84 H residues; PfLDH (316AA) has 6 R, 26 K, and 9 H residues. Given the high proportion of H residues in PfHRP2 (34.1%) and PfHRP3 (30.5%), we hypothesized that this amino acid bias might hinder trypsin accessibility. We undertook an analysis of the amino acid frequencies of PfHRP2, PfHRP3, and PfLDH and compared to the average amino acid frequencies of all 265 *P. falciparum* proteins identified in the 29 plasma samples. The amino acid distribution in PfLDH was found to align more closely with that of the 265 unique *P. falciparum* proteins detected by LC-MS proteomics. In contrast, PfHRP2 and PfHRP3 were rich in H and alanine (A), and depleted in glutamate (E), isoleucine (I), proline (P), glutamine (Q), and R (**Figure 3**), indicating possible reduced accessibility for trypsin cleavage.

**Figure 3.**
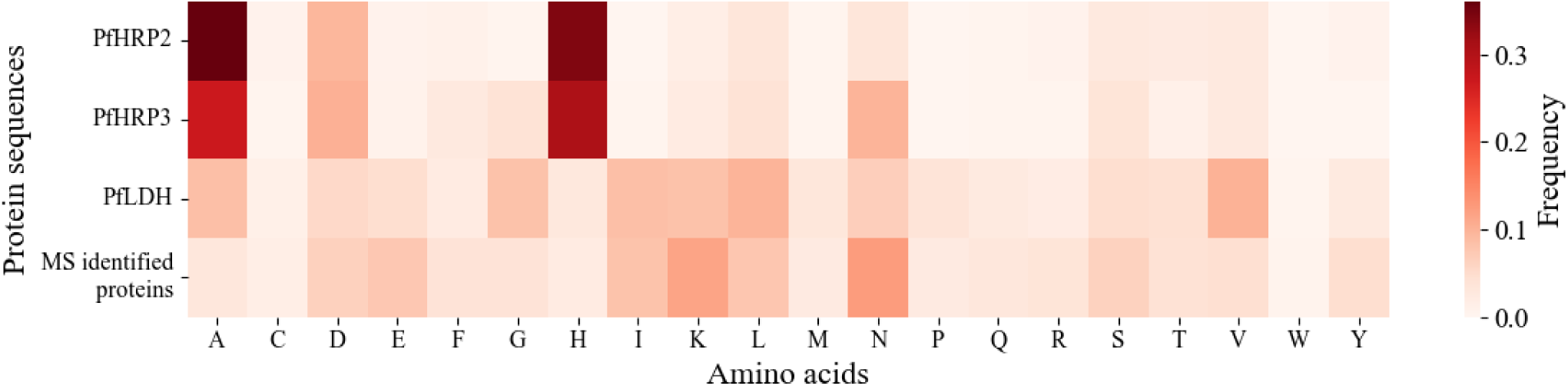
Amino acid composition of PfHRP2, PfHRP3, PfLDH, and experimentally identified *P. falciparum* proteins. Amino acids are indicated by their one letter code. **ALT Text:** Bar chart of amino acid composition for PfHRP2, PfHRP3, PfLDH compared to LC-MS identified proteins. Each bar represents the frequency of individual amino acids.

## DISCUSSION

The failure of RDTs targeting PfHRP2 to detect malaria parasites in peripheral blood is of significant concern in many African and Asian endemic countries. This study presents a molecular, transcriptomic, and proteomic characterization of *Plasmodium* infections that failed RDT detection in Mizan Aman, Ethiopia.

The results of our study demonstrate the complexity behind RDT failure and suggest that a multitude of factors can produce a false-negative diagnosis. Using molecular detection by species-specific PCR as the gold standard, the RDT we used correctly identified only 10.3% (3/29) of *P. falciparum* infections, 85.5% (53/62) of *P. vivax* infections, and 7.7% (1/13) of mixed *P. falciparum/P. vivax* infections (**Figure 1**). In *P. falciparum* PCR-confirmed samples we found significantly lower parasite densities in Pan-P.f- (i.e., RDT-) samples compared to Pan+P.f- samples (adjusted *p* = 0.006) (**Figure 2**). For *P. vivax*, higher parasitemias were strongly associated with Pan+P.f- results (*p* = 0.0001). Overall, the RDT used in our study failed to detect low parasitemia infections of either *P. falciparum* or *P. vivax*. In mixed species infections, *P. falciparum* parasite densities were consistently lower than *P. vivax*, which may further reduce the ability to detect PfHRP2 in such infections, leading to species misclassifications.

We detected dual *pfhrp2*/*pfhrp3* gene deletions in 90.9% (n=20 of 22 that passed QC) of PCR-confirmed *P. falciparum* samples; no isolates had a deletion in only one of the two genes (**Table 2**). Infections with deletion-harboring parasites had a range of parasitemias, with many of the high parasitemic samples also having gene deletions. We identified transcripts of *pfhrp2* in 28% (n=8) and of *pfhrp3* in 24% (n=7) of the 29 PCR-confirmed *P. falciparum* isolates; the 3 participants that tested PfHRP2 RDT+ all had transcripts from both genes. Interestingly, deletions in *pfhrp2* and *pfhrp3* were visible in several (n=9) isolates with transcripts from both genes. Many more of the 29 PCR-confirmed *P. falciparum* isolates had *pfldh* transcripts (n=18), although only 9/18 of these also had a positive panLDH RDT result. This high prevalence of *pfhrp2*/*pfhrp3* gene deletions aligns with recent studies by the Ethiopia Public Health Institute and others across the country that determined 10-22% of *P. falciparum* infections were missed due to the presence of deletions [13, 32]. Other groups conducting surveillance in smaller regions of the country found deletion prevalence ranging from 58-64% [15, 33]. These findings prompted release of a policy brief by the Ethiopian Ministry of Health in September 2022 where the shift to a non-PfHRP2 RDT was recommended [34].

Success of several of the assays used in our study depended upon a higher parasitemia in the isolates tested. For example, identifying *pfhrp2* and *pfhrp3* deletions in infections with the seven lowest parasitemias was not possible due to failure of the control for the qPCR assay. Similarly, PfLDH peptides were identified in 10/29 PCR-confirmed *P. falciparum* isolates that had the highest parasitemias. PfHRP2 and PfHRP3 peptides were not detected in any of the 29 *P. falciparum* isolates, including the 3 PfHRP2 RDT+ samples. Our *in silico* trypsin digestion analysis revealed limited peptide coverage for PfHRP2 (12.79%) and PfHRP3 (14.18%) compared to PfLDH (54.43%) and other *P. falciparum* proteins (average coverage: 62.61%) (**Table 3**), possibly due to the distinct amino acid composition of PfHRP2 and PfHRP3 (**Figure 3**) which likely reduces trypsin accessibility and thus peptide detection by LC-MS. In addition, highly abundant host proteins such as hemoglobin and immunoglobulins can cause ion suppression [35], which may have been a factor in our LC-MS proteomics. These findings highlight the challenges of identifying robust assays for parasite biomarker discovery in infected patient plasma.

Our study’s limitations include a smaller sample size than other published *pfhrp2/3* deletion studies conducted in Ethiopia [13, 15, 32], and the cross-sectional nature of the study. Antigen dynamics vary over the course of an infection, and *pfhrp2*/*pfhrp3* and *pfldh* gene expression and transcript abundance change as an infection progresses, after drug treatment, and depending upon the assay being used and the time point it is employed [36]. Mixed infections further complicate interpretation across detection methods [37]. Future work as part of our ongoing surveillance studies in Ethiopia will include assessment of antigen dynamics such as time-to-antigen-clearance and time-to-RDT-negativity in longitudinal cohorts [38].

In conclusion, we used a combination of species-specific PCR, gene-targeted qPCR, RNA-seq, and proteomics to characterize the *Plasmodium* parasites infecting individuals at three clinics in southwest Ethiopia. Our data indicate that RDT failure in Ethiopia is multifactorial, arising from an interplay of *pfhrp2/3* deletions, low parasite densities, and antigen dynamics that is further complicated by a prevalence of mixed infections. New parasite biomarkers that can be developed into next generation RDTs are urgently required.

## Supplementary Data

Supplementary materials are available at The Journal of Infectious Diseases online (http://jid.oxfordjournals.org/). Supplementary materials consist of data provided by the author that are published to benefit the reader. The posted materials are not copyedited. The contents of all supplementary data are the sole responsibility of the authors. Questions or messages regarding errors should be addressed to the author.

## Data Availability

All data produced in the present study are available upon reasonable request to the authors

## Acknowledgments

We are indebted to the communities served by Mizan Teaching Hospital, Mizan Health Center, and Kite Health Center in the Mizan Teferi municipality and the teams that staff these facilities.

## Disclaimer

The study funders had no role in the study design, implementation, analysis, manuscript preparation, or decision to submit this article for publication.

## Financial support

Research reported in this publication was supported by the National Institute of Allergy and Infectious Diseases of the National Institutes of Health under Award Number U19AI089676 (JC) and by Bloomberg Philanthropies as part of their support of the Johns Hopkins Malaria Research Institute. The content is solely the responsibility of the authors and does not necessarily represent the official views of the National Institutes of Health or other funders. MK and HS were partially supported by a New York University MacCracken fellowship.

## Potential conflicts of interest

All authors: No reported conflicts of interest. All authors have submitted the ICMJE Form for Disclosure of Potential Conflicts of Interest. Conflicts that the editors consider relevant to the content of the manuscript have been disclosed.

## Author contributions

JC, GY, and DY conceived and obtained funding for the study. MN, DL, and JC designed the study. TT, TTe, DT, BL, DL, MZ, and FL collected the samples and clinical data at the health centers. MN and AP processed the samples, and AK provided assay training and guidance. ZW and HS undertook the proteomics sample prep and data analysis. MN, AK, and JC analyzed the data. MN, AK, ZW, HS, SS, and JC wrote and edited the paper. All authors have read and approved the final manuscript.

## Notes

### Competing Interest Statement

The authors have declared no competing interest.

### Author Declarations

The study protocols were approved by the National Research and Ethics Review Committee (NRERC), Ministry of Education, and the Federal Democratic Republic of Ethiopia (Reference No. 03/246/9S/22).

