## Supplementary Table 1 for "Molecular, Transcriptomic, and Proteomic Characterization of *Plasmodium* Infections that Evade Detection by Rapid Diagnostic Tests in Mizan Aman, Ethiopia"

**Supplementary Table 1.** qPCR assay primers used for identification of *pfhrp2* and *pfhrp3* gene deletions, with *pfldh* and human beta-tubulin (*HumTuBB*) genes used as controls.

**ALT Text:** Table showing qPCR assay primers for detecting *pfhrp2* and *pfhrp3* deletions, with *pfldh* and human beta-tubulin genes as controls. Table includes primer names, sequences, and target genes.

| **Primer/Probe** | **Sequence** | **Primer Specificity** |
| --- | --- | --- |
| Pfhrp2_F1 | 5'-TAATTSCGYATTTAATAATAACTTGTG-3' | ***P. falciparum hrp2*** |
| Pfhrp2_R2 | 5'- CATCATCTACATGTGCTGGAG-3' |  |
| Pfhrp2_probe (495-520) | **FAM**-ATGCAAAAGGACTTAATTTAAATAAGAGATT-**BHQ2** |  |
| Pfhrp3_F2 | 5'- TCCGAATTTAACAATAACTTGTTTAGC-3' | ***P. falciparum hrp3*** |
| Pfhrp3_R2 | 5'- GTCAAGCACATGCAGGTGATG-3' |  |
| Pfhrp3_probe (529-555) | **JOE**-ATGCAAAAGGACTTAATTCAAATAAGAGATTA-**BHQ1** |  |
| Pfldh_F | 5'- ACGATTTGGCTGGAGCAGAT -3' | ***P. falciparum***  ***ldh*** |
| Pfldh_R | 5'- TCTCTATTCCATTCTTTGTCACTCTTTC -3' |  |
| Pfldh_probe (575-608) | **ROX**-GTAATAGTAACAGCTGGATTTACCAAGGCCCCA-**BHQ2** |  |
| HumTuBB_F | 5'- AAGGAGGTCGATGAGCAGAT -3' | **Human**  **beta-tubulin** |
| HumTuBB_R | 5'- GCTGTCTTGACATTGTTGGG -3' |  |
| HumanTuBB_P (648-668) | **CY5**-TTAACGTGCAGAACAAGAACAGCAGCT-**BHQ2** |  |
